# Secondary traumatic stress and burnout in healthcare workers during COVID-19 outbreak

**DOI:** 10.1101/2020.09.13.20186692

**Authors:** F. Marzetti, G. Vagheggini, C. Conversano, M. Miccoli, A. Gemignani, R. Ciacchini, E. Panait, G. Orrù

## Abstract

**Aims:** To assess the level of professional burnout and secondary traumatic stress, and to identify potential risk or protective factors among health care workers (HCWs) during the coronavirus disease 2019 (COVID-19) outbreak.

**Materials and Methods:** This cross-sectional study, based on an online survey, collected demographic data and mental distress outcomes from 184 HCWs from May 1st, 2020, to June,15th, 2020, from 45 different countries. The degree of secondary traumatization was assessed using the Secondary Traumatic Stress Scale (STSS), the degrees of perceived stress and burnout were assessed with Perceived Stress Scale (PSS) and Maslach Burnout Inventory Human Service Survey (MBI-HSS) respectively. Stepwise multiple regression analysis was performed to identify potential risk and protective factors for STS.

**Results:** 184 HCWs (M=90; Age mean: 46.45; SD:11.02) completed the survey. A considerable proportion of HCWs had symptoms of secondary traumatic stress (41.3%), emotional exhaustion (56.0%), and depersonalization (48.9%). The prevalence of secondary traumatic stress in frontline HCWs was 47.5% while in HCWs working in other units it was 30.3% (p<.023); additionally, the prevalence of the same outcome was 67.1% for the HCWs exposed to patients’ death and 32.9% for those HCWs which were not exposed to the same condition (p<.001). In stepwise multiple regression analysis, perceived stress, emotional exhaustion and exposure to patients’ death remained as significant predictors in the final model for secondary traumatic stress (adjusted R2 = 0.537, p< 0.001).

**Conclusions:** During the current COVID-19 pandemic, HCWs facing patients’ physical pain, psychological suffering, and death are more likely to develop secondary traumatization.

## INTRODUCTION

The health emergency due to the COVID-19 outbreak has heavily impacted the psychological and emotional wellbeing of general population^1,2^ and healthcare workers (HCWs). In frontline HCWs, different sources of psychological distress have been reported, such as uncertainty to the disease progression (short and long term effects), and treatment, lack of personal protective equipment (PPE), physical exhaustion and overwhelming workload, concerns about the direct exposure to COVID-19 at work: in particular, the latter is associated to the fear of getting infected or spreading the infection among colleagues and families members^3–8^. Additionally, frontline HCWs took care of patients who were both physically and psychologically suffering for the emergency situation (*vicarious traumatization*), and, as a consequence, they’re exposed at the risk of developing secondary traumatic stress disorder ^9,10^.

Results emerging from empirical researches, carried out in comparable periods such as the Severe Acute Respiratory Syndrome (SARS) or the Middle East Respiratory Syndrome (MERS) outbreaks, highlighted that HCWs experienced high levels of stress, anxiety and depressive symptoms ^11,12^, psychological distress ^13^ and post-traumatic stress symptoms that include avoidance, hyperarousal, and insomnia^11,14,15^. As expected, frontline HCWs experienced a greater psychological distress compare to HCWs with secondary roles ^11^.

According to recent studies, Chinese HCWs directly caring for COVID-19 patients showed higher levels of distress, anxiety and insomnia while compared to HCWs in secondary roles ^16–19^. Medical HCWs showed a higher prevalence of insomnia, depressive, anxiety and obsessive-compulsive symptoms compared to nonmedical healthcare workers ^18^. Two recent systematic reviews and meta-analysis underlined the higher prevalence of depression, anxiety, and insomnia among HCWs during the COVID-19 outbreak ^20,21^.

Direct exposure to high level of distress during COVID-19 pandemic, seems to increase the risk for long term consequences such as post-traumatic stress, depressive symptoms ^22^ or professional burnout with adverse outcomes for the whole organization ^23^. Professional burnout or occupational burnout has been defined by Maslach ^24^ as a *“response to prolonged and chronic stress at the workplace, characterized by three dimensions: emotional exhaustion, depersonalization and reduction of personal abilities”*. This condition seems to be predominant within the medical health care professionals than others ^25,26^. Schanafelt et al.^27^ reported that the overall mean rate of physician burnout rose from 45.5% in 2011 to 54.4% in 2014 (p<.001). It is characterized by a gradual development over time accompanied by reduced professional satisfaction which can lead to a worse ability to judge, late or inadequate responses to changes in the clinical context, and lack of patient confidence in HCWs, compromising professional performance^26,28–31^.

Another consequence of the COVID-19 outbreak may be represented by pathologic secondary traumatic stress (STS). Figley^32^ defined STS as *“the stress deriving from helping others who are suffering or who have been traumatized”*. Some authors use the term STS or compassion fatigue or vicarious traumatization interchangeably ^33^. In ordinary situations, because of the implications of their professional sector, HCWs may be at higher risk of developing pathological secondary traumatization and this is particularly true now more than before, considering the present emergency situation. Protective factors such as resilience, self-efficacy and perceived social support may be able to reduce STS and anxiety symptoms^34^.

Interventions aimed at healthcare professionals should, through the support of psychologists, focusing on the management and containment of maladaptive behaviors and broader range of emotional disorders in order to reduce stress and improve professional performance ^17,35–37^. Even if different kind of psychological interventions have been released, results remain unclear and HCWs often refuse to participate ^17,38,39^.

The aim of the present study is to assess the psychological distress in terms of perceived stress, professional burnout and STS, and to identify potential risk or protective factors, among HCWs during the COVID-19 outbreak all over the world.

## MATERIALS AND METHODS

### Study design and participants

A cross-sectional international survey addressed to HCWs was conducted from May 1 to June 15, 2020. The Ethics Committee of the University of Pisa approved our study survey and procedures of informed consent before the formal survey.

A link to the above-mentioned survey was sent directly to HCWs and to the European Respiratory Society (ERS) and was published at the following link: https://www.ersnet.org/research/covid-19-surveys. The informed consent was available online and was provided to all participants before enrolment. The survey was anonymous and confidentiality of information was assured.

One-hundred and eighty-four HCWs from 45 Countries and 5 continents completed the online survey. Participants were eligible if the following criteria were met: 1. working in health care during COVID-19 outbreak; 2. give informed consent.

### Materials

Socio-demographic data were self-reported by participants and included gender, age, Country, education, occupation, seniority, civil status, number of children and pathologies.

Data related to personal and professional experience during COVID-19 outbreak were also included: actual lockdown policies of the belonging Country, positivity for COVID-19, severity of symptoms of family members or friends infected by COVID-19, direct involvement in the assistance of COVID-19 patients, daily working with COVID-19 patients, exposure to patients’ death. Respondents were also asked to evaluate how the emergency situation was managed by the organization/hospital (1 very bad management – 10 very good management) and the perceived degree of emergency (1 COVID-19 is not a real emergency – 10 it’s a real emergency).

#### Perceived Stress Scale (PSS)

The PSS is a 10-item questionnaire designed to assess the degree to which external demands seems to exceed the individual perceived ability to cope^40^. Respondents are asked to indicate how frequently they felt or thought certain way in the last month on a 5-point Likert scale ranging from 0 (never) to 4 (very often). The PSS total score is calculated by summing up the item scores, with a higher score indicating higher perceived stress. Score range between 0 to 40.

#### Secondary Traumatic Stress Scale (STSS)

The STSS is a 17-item questionnaire designed for measure the negative impact of indirect exposure to traumatic events in HCWs caring for suffering or traumatized clients. The traumatic stressor for HCWs is identified as exposure to patients. Respondents are asked to indicate how frequently the item was true for them in the past seven days on a 5-point Likert scale ranging from 1 (never) to 5 (very often). The STSS has a global score and three subscales: Intrusion (five items), that refers to recurrent and intrusive distressing recollections of patients, including images, thoughts or perceptions; Avoidance (seven items), that measures the avoidance of stimuli associated with the care of patients and the numbing of general responsiveness; Arousal (five items), that assess symptoms such as irritability, hypervigilance, difficulty concentrating, etc… The STSS global score is calculated by summing up all the item scores, with a higher score indicating a higher frequency of symptoms. The total score ranges from 17 to 85 and is categorized into no/little (17-28), mild (28-37), moderate (38-43), high (44-48), and severe (49-85) levels of secondary trauma ^41^.

#### Maslach Burnout Inventory Human Service Survey (MBI-HSS)

The MBI-HSS is a 22-item questionnaire that assesses professional burnout within people involved in the care and social services ^24,42^. Respondents are asked to indicate the frequency with which they experience certain feelings or attitudes on a 7-point Likert scale ranging from 0 (never) to 6 (every day). MBI-HSS is composed by three subscales: Emotional Exhaustion (MBI-EE, 9 items), that assess the feelings of being emotionally overextended by one’s work; Depersonalization (MBI-D, 5 items), that measures unfeeling and impersonal response to care; Personal Accomplishment (MBI-PA, 8 items), that assess the feelings of competence, the perceived effectiveness on the job. The scores for each subscale are not combined into a global score: they are separated, with different cut-offs points (MBI-EE: low 0-16, moderate 17-26, high 27-54; MBI-D: low 0-6, moderate 7-12, high 13-35; MBI-PA: low 0-31, moderate 32-38, high 39-48).

#### 14-Item Resilience Scale (RS-14)

The RS-14 is a 14-item questionnaire to assess the individual ability to withstand or adaptively recover from stressor ^43^. Items are evaluated on a 7-point Likert scale ranging from 1 (strongly disagree) to 7 (strongly agree). Total score ranges from 14 to 98, with higher scores indicating greater resilience. RS-14 yields reliable scores, coefficient alphas of .90 and greater ^43^.

#### General Self-Efficacy Scale (GSE)

The GSE is a 10-items instrument that measures the perceived self-efficacy, the belief that one can successfully cope with adverse situation or stressor ^44^. GSE explicitly refers to personal agency, that is, the belief that the own actions are responsible for successful outcomes. Each item is evaluated on a 4-point Likert scale, scored from 1 (not at all true) to 4 (completely true). The total score is calculated by finding the sum of all items and ranges between 10 and 40, with a higher score indicating higher self-efficacy.

### Statistical Analysis

Data are presented as mean with standard deviation (SD). Comparisons between groups were performed using t-test for independent samples and chi-squared test for categorical variables. Pearson’s correlation was computed to evaluate the relationship between psychological distress variables, protective factors, socio-demographic characteristics, and COVID experience. Stepwise multiple linear regression analysis was performed to determine potential risk and protective factors for secondary traumatic stress. P values <0.05 were considered statistically significant. The analysis was conducted using IBM SPSS statistics version 21.

## RESULTS

### Demographic characteristics and outcomes of psychological distress in total cohort

A total of 184 HCWs (M=90; Age mean: 46.45; SD:11.02) completed the survey questionnaire a Demographic characteristics and outcomes of psychological distress in totalnd the professions were the following: physicians (n=138; 75.0%), nurses (n=10; 5.4%), surgeons (n=3; 1.6%), psychologists (n=2; 1.1%) and other health professionals (n=31; 16.8%). The demographic characteristics of the sample and the Country of origin are summarized in **Table 1 and Table 2**, respectively.

118 HCWs (64.1%) were frontline and directly involved in the care of COVID-19 patients while 66 HCWs (35.9%) were involved in different units. Ten out of 184 HCWs (5.6%) were infected by COVID-19 and 57 HCWs (31.0%) had one or more family members infected by COVID-19. The mean of respondents was mainly satisfied with how the organization managed the critical situation (mean=7.73, SD=1.75) and did not perceive COVID-19 outbreak as a severe emergency (mean=4.28, SD=3.15) (**Table 3**).

**Table 1:**
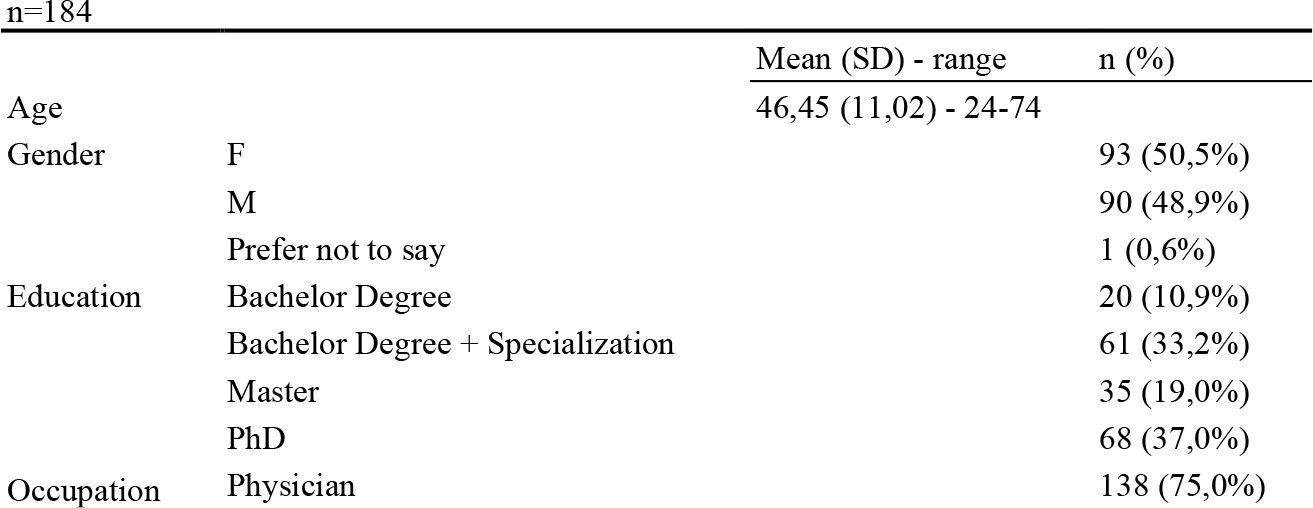
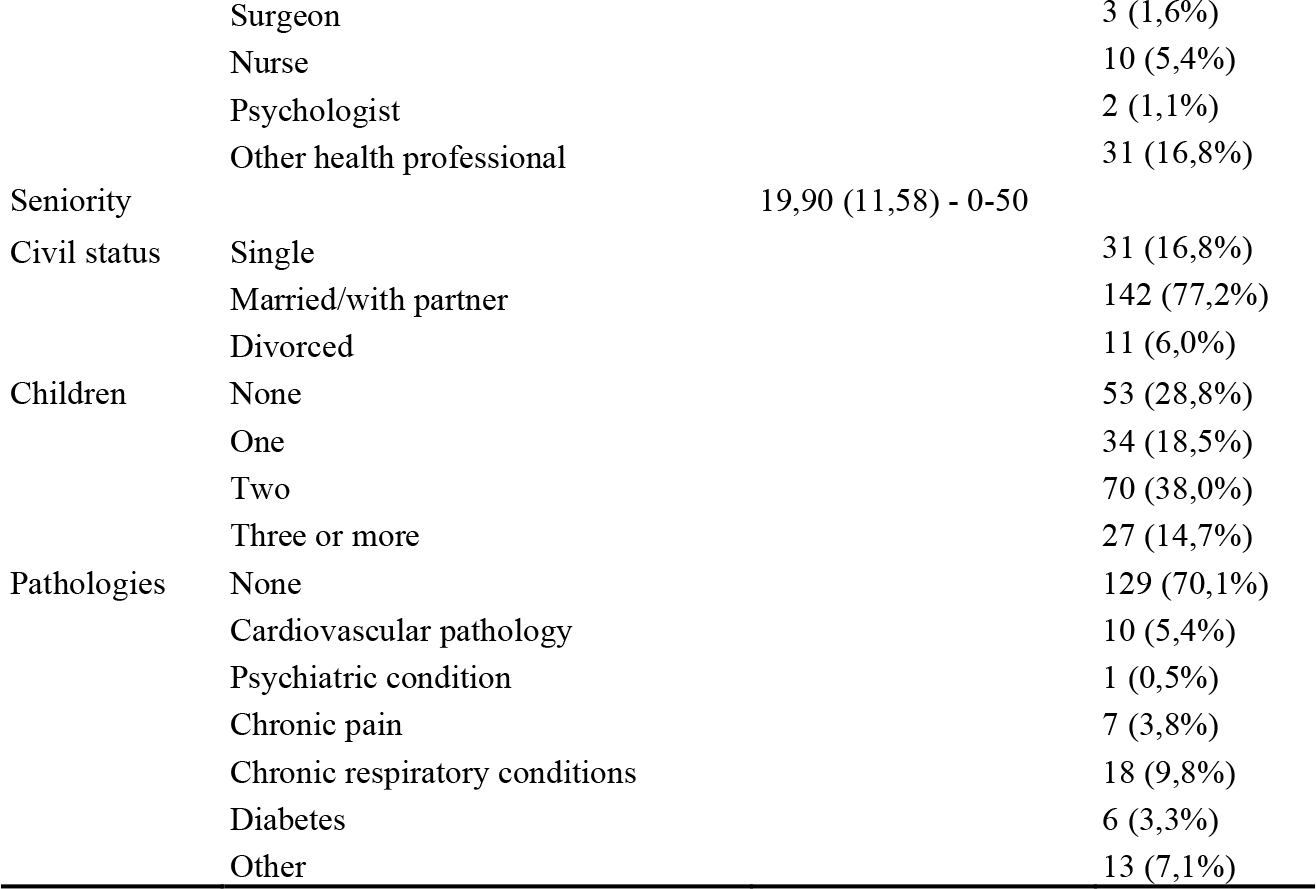
Demographic characteristics

**Table 2:** Distribution per Country

| n=184 |  |  |  |  |  |  |  |  |  |
| --- | --- | --- | --- | --- | --- | --- | --- | --- | --- |
|  | n | % | Lock-down policies |  |  |  | n | % | Lock-down policies |
| Algiers | 1 | 0,5 | % | Hard | Latvia | 1 | 0,5 | % | Hard |
| Argentina | 1 | 0,5 | % | Hard | Lebanon | 1 | 0,5 | % | Soft |
| Australia | 1 | 0,5 | % | Soft | Macedonia | 1 | 0,5 | % | - |
| Austria | 2 | 1,1 | % | Hard | Malaysia | 4 | 2,2 | % | Hard |
| Belgium | 3 | 1,6 | % | Soft | Mexico | 1 | 0,5 | % | Soft |
| Brazil | 8 | 4,3 | % | Soft | Netherlands | 10 | 5,4 | % | Hard |
| Canada | 2 | 1,1 | % | Soft | Philippines | 2 | 1,1 | % | Soft |
| Chile | 1 | 0,5 | % | Hard | Poland | 1 | 0,5 | % | Soft |
| Colombia | 2 | 1,1 | % | Hard | Portugal | 6 | 3,3 | % | Hard |
| Croatia | 4 | 2,2 | % | Hard | Romania | 3 | 1,6 | % | Soft |
| Czech Republic | 2 | 1,1 | % | Hard | Serbia | 3 | 1,6 | % | Soft |
| Denmark | 2 | 1,1 | % | Soft | South Africa | 1 | 0,5 | % | Hard |
| Estonia | 1 | 0,5 | % | Soft | Spain | 4 | 2,2 | % | Hard |
| Finland | 1 | 0,5 | % | Soft | Sri Lanka | 1 | 0,5 | % | Soft |
| France | 4 | 2,2 | % | Hard | Sweden | 3 | 1,6 | % | Soft |
| Germany | 5 | 2,7 | % | Hard | Switzerland | 2 | 1,1 | % | Hard |
| Greece | 3 | 1,6 | % | Hard | Thailand | 1 | 0,5 | % | Soft |
| Hungary | 1 | 0,5 | % | Soft | Turkey | 2 | 1,1 | % | Hard |
| India | 3 | 1,6 | % | Hard | Ucraina | 1 | 0,5 | % | Soft |
| Indonesia | 9 | 4,9 | % | Soft | UK | 23 | 12,5 | % | Soft |
| Ireland | 7 | 3,8 | % | Soft | Uruguay | 1 | 0,5 | % | - |
| Italy | 39 | 21,2 | % | Hard | USA | 3 | 1,6 | % | Soft |
| Korea | 1 | 0,5 | % | Hard | missing | 6 | 3,3 | % | - |

Note: Lockdown policies were classified as hard if a complete lockdown was applied or soft if only partial limitations were established<sup>47</sup>.

**Table 3:**
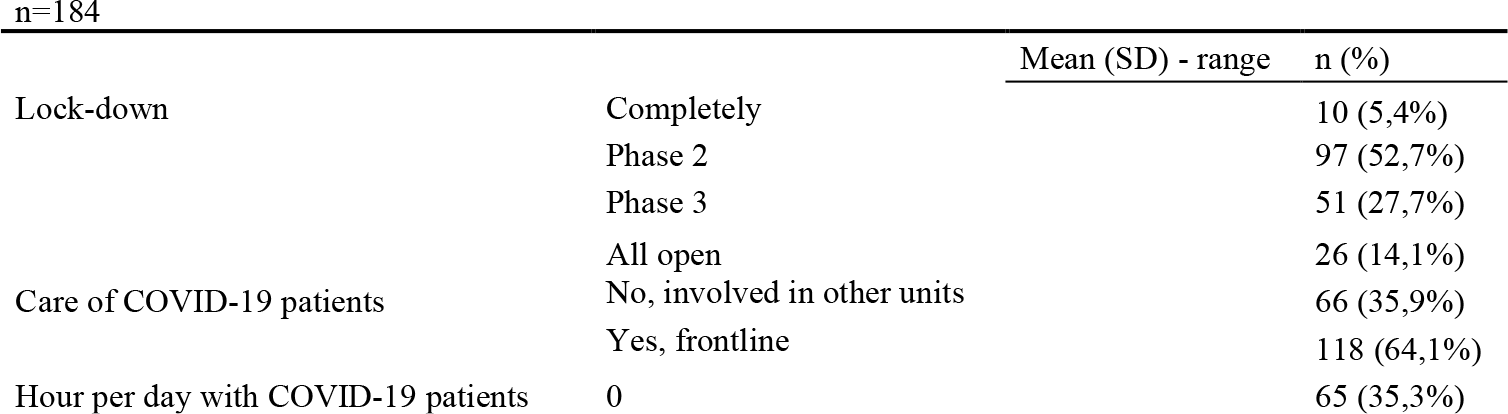
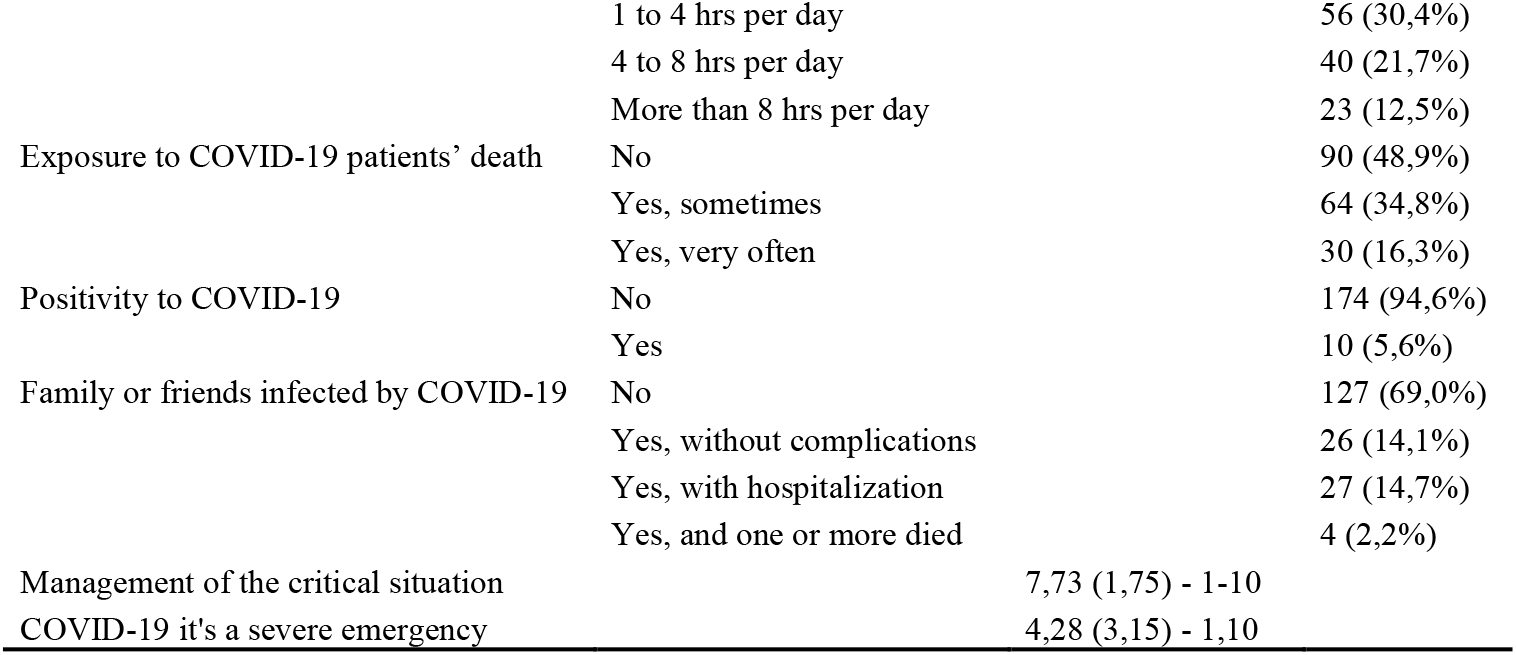
COVID-19 outbreak individual experience

A considerable proportion of HCWs had symptoms of secondary traumatic stress (STSS≥38, moderate to severe symptoms, 41.3%), emotional exhaustion (MBI-EE≥17, moderate to high, 56.0%), and depersonalization (MBI-D≥7, moderate to high, 48.9%). The mean (SD) scores on the PSS, STSS and subscales, MBI-HSS subscales, GSE, and RS-14 are shown in **Table 4**.

**Table 4:** Questionnaire scales and subscales scores and prevalence in total cohort

| n=184 |  |  |
| --- | --- | --- |
|  | Mean (SD) | n (%) |
| <u>PSS</u> | 16,80 (6,27) |  |
| <u>STSS</u> | 36,41 (12,79) |  |
| No STSS |  | 65 (35,4%) |
| Mild STSS |  | 43 (23,4%) |
| Moderate STSS |  | 26 (14,1%) |
| High STSS |  | 8 (4,3%) |
| Severe STSS |  | 42 (22,8%) |
| Intrusion | 10,45 (4,08) |  |
| Avoidance | 14,50 (5,32) |  |
| Arousal | 11,46 (4,50) |  |
| <u>MBI-HSS</u> |  |  |
| EE | 19,66 (11,12) |  |
| Low |  | 81 (44,0%) |
| Moderate |  | 56 (30,5%) |
| High |  | 47 (25,5%) |
| D | 7,53 (5,51) |  |
| Low |  | 94 (51,1%) |
| Moderate |  | 58 (31,5%) |
| High |  | 32 (17,4%) |
| PA | 32,48 (8,16) |  |
| Low |  | 70 (38,0%) |
| Moderate |  | 70 (38,0%) |
| High |  | 44 (24,0%) |
| <u>GSE</u> | 30,75 (5,73) |  |
| <u>RS-14</u> | 78,61 (12,71) |  |

Abbreviations: PSS=Perceived Stress Scale; STSS=Secondary Traumatic Stress Scale; MBI-HSS=Maslach Burnout Inventory Health Services Scale; EE= Emotional Exhaustion; D=Depersonalization; PA= Personal Accomplishment; GSE=General Self-Efficacy Scale; RS-14=14-item Resilience Scale.

Correlation analysis among psychological distress, secondary traumatic stress, professional burnout, protective factors, demographics, and professional experience during COVID-19 outbreak is reported in **Table 5**.

**Table 5:** Significant Pearson correlation coefficients r: association between perceived stress, secondary traumatic stress, professional burnout, protective factors, demographics and COVID-19 individual experience in total cohort (n=184)

|  | 1 | 2 | 3 | 4 | 5 | 6 | 7 | 8 | 9 | 10 | 11 | 12 | 13 | 14 | 15 | 16 |
| --- | --- | --- | --- | --- | --- | --- | --- | --- | --- | --- | --- | --- | --- | --- | --- | --- |
| 1.PSS | - |  |  |  |  |  |  |  |  |  |  |  |  |  |  |  |
| 2.STSS_ | .633** | - |  |  |  |  |  |  |  |  |  |  |  |  |  |  |
| 3.Intrusion | .540** | .893** | - |  |  |  |  |  |  |  |  |  |  |  |  |  |
| 4.Avoidance | .591** | .939** | .752** | - |  |  |  |  |  |  |  |  |  |  |  |  |
| 5.Arousal | .611** | .923** | .742** | .805** | - |  |  |  |  |  |  |  |  |  |  |  |
| 6.MBI-EE | .555** | .645** | .562** | .596** | .621** | - |  |  |  |  |  |  |  |  |  |  |
| 7.MBI-D | .416** | .480** | .443** | .472** | .404** | .586** | - |  |  |  |  |  |  |  |  |  |
| 8.MBI-PA | -.307** | -.034 | -.018 | -.090 | -.006 | -.095 | -.192** | - |  |  |  |  |  |  |  |  |
| 9.GSE | -.381** | -.244** | -.214** | -.268** | -.182* | -.259** | -.190** | .466** | - |  |  |  |  |  |  |  |
| 10.RS-14 | **-.337\*\*** | **-.293\*\*** | **-.234\*\*** | **-.300\*\*** | **-.264\*\*** | **-.374\*\*** | **-.341\*\*** | **.498\*\*** | **.565\*\*** | - |  |  |  |  |  |  |
| 11.Age | **-.321\*\*** | **-.160\*** | -.135 | -.143 | **-.165\*** | -.131 | **-.148\*** | **.202\*\*** | .130 | .121 | - |  |  |  |  |  |
| 12.Gender | .123 | .139 | **.183\*** | .078 | .138 | **.199\*\*** | -.020 | .067 | -.136 | -.049 | **-.262\*\*** | - |  |  |  |  |
| 13.Children | **-.313\*\*** | **-.302\*\*** | **-.268\*\*** | **-.284\*\*** | **-.280\*\*** | **-.259\*\*** | -.122 | **.160\*\*** | **.158\*** | .143 | **.435\*\*** | **-.321\*\*** | - |  |  |  |
| 14.C19_ff | **.201\*\*** | **.146\*** | **.185\*** | .072 | **.163\*** | .055 | .027 | .065 | .034 | .059 | -.020 | .138 | .006 | - |  |  |
| 15.C19_hour per day | **.177\*** | **.206\*\*** | **.246\*\*** | **.161\*** | **.173\*** | .037 | -.009 | .022 | -.021 | .048 | -.083 | -.092 | -.047 | .093 | - |  |
| 16.C19_death | **.197\*\*** | **.261\*\*** | **.304\*\*** | **.180\*\*** | **.254\*\*** | .110 | -.016 | .130 | .091 | .103 | -.078 | -.074 | .001 | **.215\*\*** | **.671\*\*** | - |

Abbreviations: PSS=Perceived Stress Scale; STSS=Secondary Traumatic Stress Scale; MBI-EE=Maslach Burnout Inventory Health Services Scale, Emotional Exhaustion; MBI-D=Maslach Burnout Inventory Health Services Scale, Depersonalization; MBI-PA= Maslach Burnout Inventory Health Services Scale, Personal Accomplishment; GSE=General Self-Efficacy Scale; RS-14=14-item Resilience Scale; C19\_ff=Family members or friends infected by COVID19; C19\_hour per day= Hour per day with COVID-19 patients; C19\_death=Exposed to patients with COVID-19 death. Bold data indicate correlations that are significant. \*p < .05; \*\* p < .01.

### Demographic characteristics and outcomes of psychological distress: differences between subgroups

Women HCWs showed significant higher scores than men HCWs on STSS Intrusion subscale (p=.013), and on MBI-EE (p=.007). HCWs without children exhibited significant higher scores on global STSS (p<.001) and all subscales (Intrusion, p=.003; Avoidance, p<.001; Arousal, p=.001), PSS (p=.001), MBI-EE (p=.002) and MBI-D (p=.033), compared to the colleagues with one or more children, and lower GSE (p=.031). HCWs with family members or friends infected by COVID-19 displayed significant higher scores on PSS (p=.013), Intrusion (p=.028) and Arousal subscale (p=.057) **(Table 6)**.

**Table 6:**
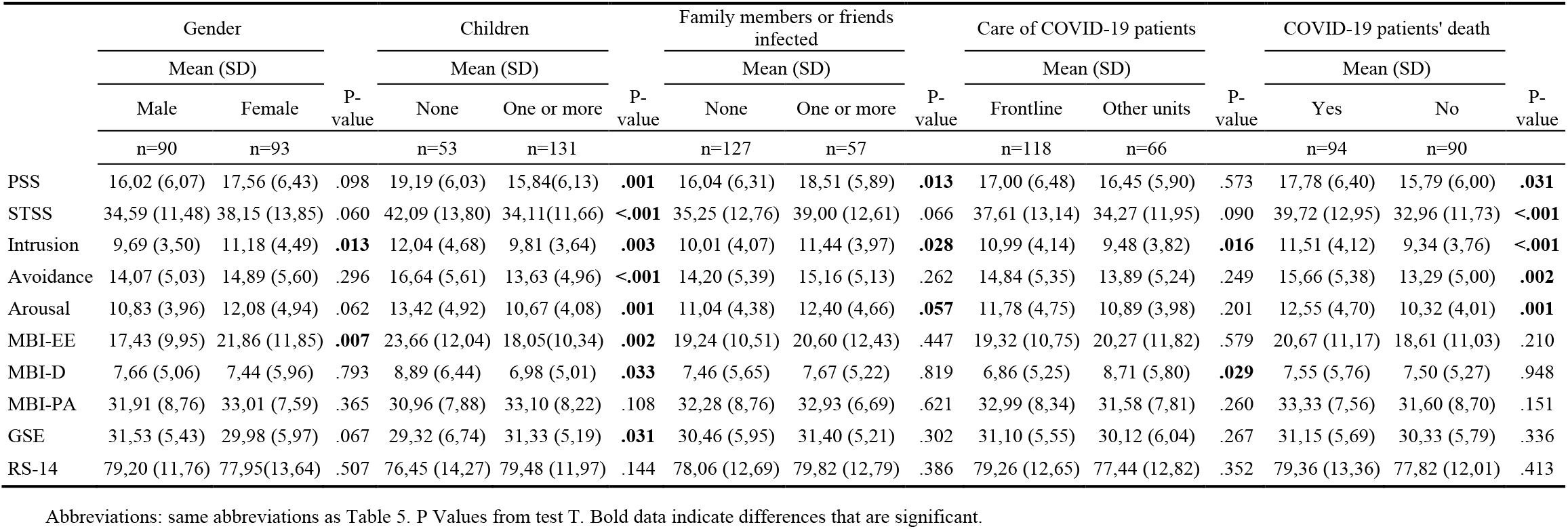
Questionnaire scales and subscales scores in subgroups

The comparison between HCWs working in Countries with hard lock-down policies (HLD, n=96) and HCWs working in Countries with softer lock-down policies (SLD, n=80) showed that HCWs working in SLD conditions exhibited lower MBI-D (p=.054) scores and higher MBI-PA (p=.019) and RS-14 (p=.005) scores.

#### Frontline HWCs

The prevalence of STS (STSS≥38, moderate to severe symptoms) in frontline HCWs (F-HCWs, n=118) was 47.5% while a lower rate (30.3%) was detected for the HCWs working in other units (OU-HCWs, n = 66) (p<.029). F-HCWs exhibited significant higher scores on STSS Intrusion subscale (p=.016) than OU-HCWs, but significant lower scores on MBI-D (p=.029).

Correlation analysis in F-HCWs subgroup showed that the higher Intrusion scores were significantly and positively associated to PSS (p<.001), MBI-EE (p<.001), MBI-D (p<.001), female gender (p=.004), hours per day spent with patients (p=.020) and exposure to patients’ deaths (p=.001). Meanwhile, they were negatively related to age (p=.003), number of children (p=.002), GSE (p=.002), RS-14 (p.036). In OU-HCWs subgroup, a positive significant correlation was found between Intrusion and PSS (p<.001), MBI-EE (p<.001), MBI-D (p<.001), while a negative relation was found with RS-14 (p=.003) and number of children (p=.021) (**Table 7**).

**Table 7:**
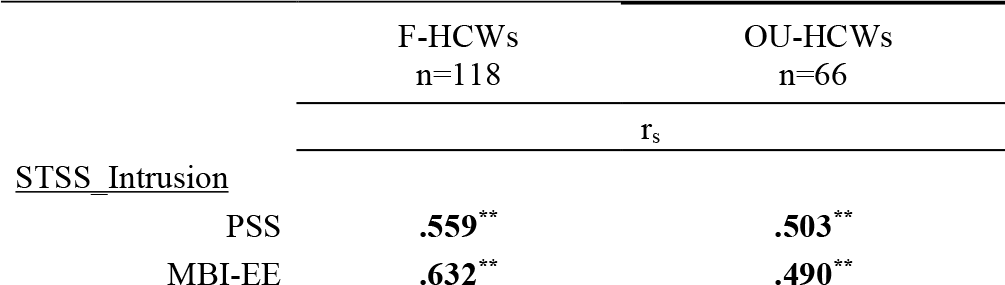
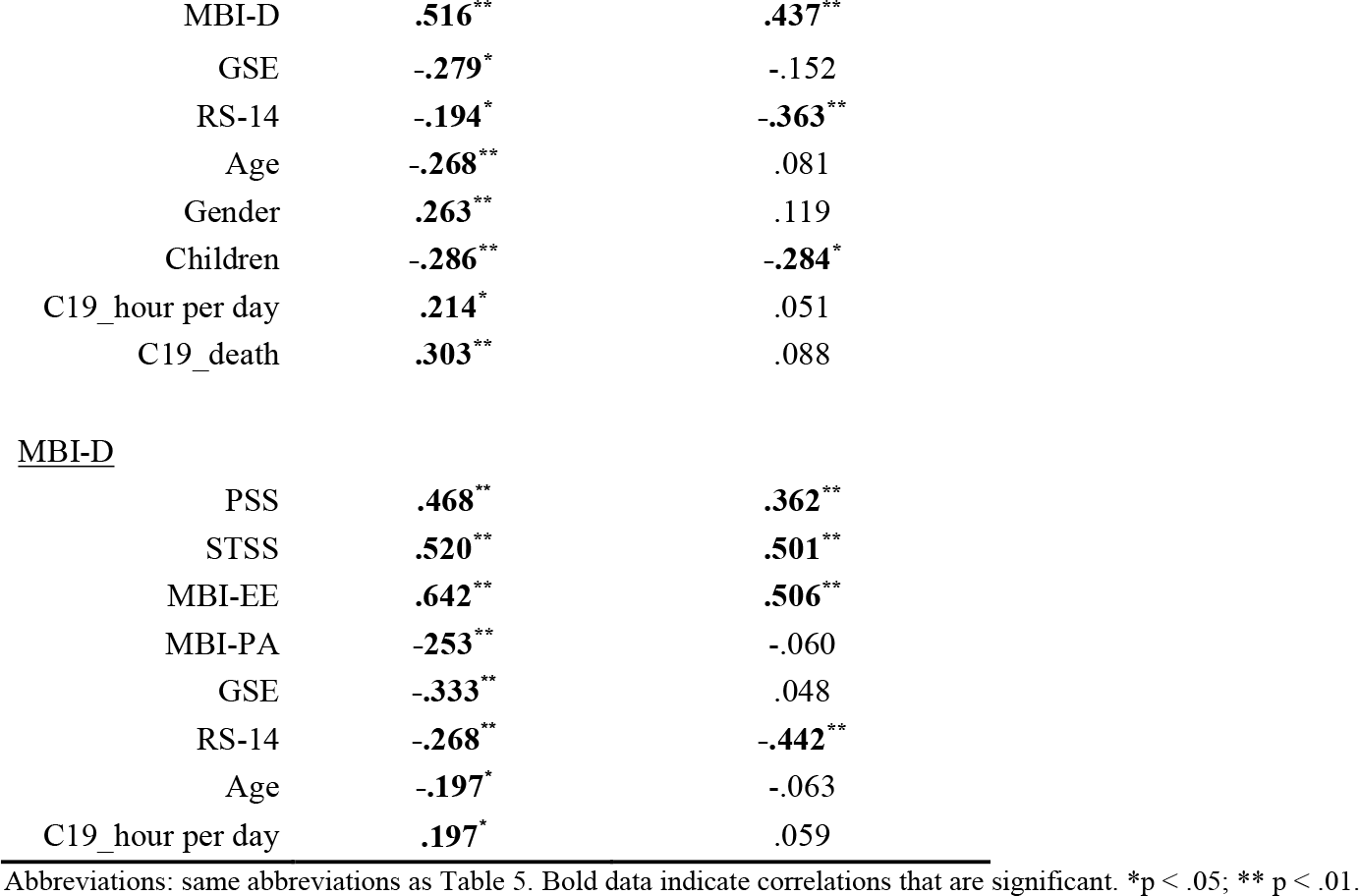
Significant Pearson correlation coefficients r: associations between Intrusion and Depersonalization, in F-HCWs vs OU-HCWs

|  | F-HCWs<br>n=118 | OU-HCWs<br>n=66 |
| --- | --- | --- |
| | $r_s$ | |
| <u>STSS_Intrusion</u> |  |  |
| PSS | .559** | .503** |
| MBI-EE | .632** | .490** |
| MBI-D | **.516\*\*** | **.437\*\*** |
| GSE | **-.279\*** | -.152 |
| RS-14 | **-.194\*** | **-.363\*\*** |
| Age | **-.268\*\*** | .081 |
| Gender | **.263\*\*** | .119 |
| Children | **-.286\*\*** | **-.284\*** |
| C19_hour per day | **.214\*** | .051 |
| C19_death | **.303\*\*** | .088 |
| MBI-D |  |  |
| PSS | **.468\*\*** | **.362\*\*** |
| STSS | **.520\*\*** | **.501\*\*** |
| MBI-EE | **.642\*\*** | **.506\*\*** |
| MBI-PA | **-.253\*\*** | -.060 |
| GSE | **-.333\*\*** | .048 |
| RS-14 | **-.268\*\*** | **-.442\*\*** |
| Age | **-.197\*** | -.063 |
| C19_hour per day | **.197\*** | .059 |

Abbreviations: same abbreviations as Table 5. Bold data indicate correlations that are significant. \*p < .05; \*\* p < .01.

The lower MBI-D scores found in F-HCWs had a significant and positive association with PSS (p<.001), STSS (p<.001), MBI-EE (p<.001) and the number of hours per day spent with patients (p=.033) and a significative negative association with age (p=.033), GSE (p<.001), RS-14 (p=.003), and MBI-PA (p=.006). In OU-HCWs a positive significant correlation was found between MBI-D and PSS (p=.003), STSS (p<.001), MBI-E (p<.001) meanwhile a negative correlation was reported with RS-14 (p<.001).

#### Exposure to patients’ death as a risk factor for secondary traumatic stress

The prevalence of secondary traumatic stress (STSS≥38, moderate to severe symptoms) in HCWs exposed to infected patients’ death (E-HCWs, n=94) was 54.3% while it was 27.8% in HCWs who were not exposed (NE-HCWs, n = 90) (p<.001). E-HCWs also reported significant higher scores on PSS (p=.031), STSS (p<.001), and all subscales (Intrusion, p<.001; Avoidance, p=.002; Arousal, p=.001) than NE-HCWs.

Stepwise multiple linear regression analysis was performed to find out the predictors of STSS in the total cohort. In the final model for STSS, exposure to patients’ deaths, PSS, and MBI-EE scores remained as significant predictors, with a good level of fit with the data (adjusted R^2^ =0.537). Significant protective factors, such as resilience or self-efficacy, were not found. With respect to F-HCWs and OU-HCWs subgroups, stepwise multiple regression analysis was performed to identify the predictors of Intrusion symptoms. In F-HCWs the final model for Intrusion had PSS, MBI-EE, MBI-D, female gender, and exposure to patients’ deaths as significant predictors, with a good level of fit with the data (adjusted R^2^ =0.486). Meanwhile, in the final regression model for Intrusion in OU-HCWs (adjusted R^2^=.306), only PSS and MBI-D remained as significant predictors. The results are summarized in **Table 8**.

**Table 8:**
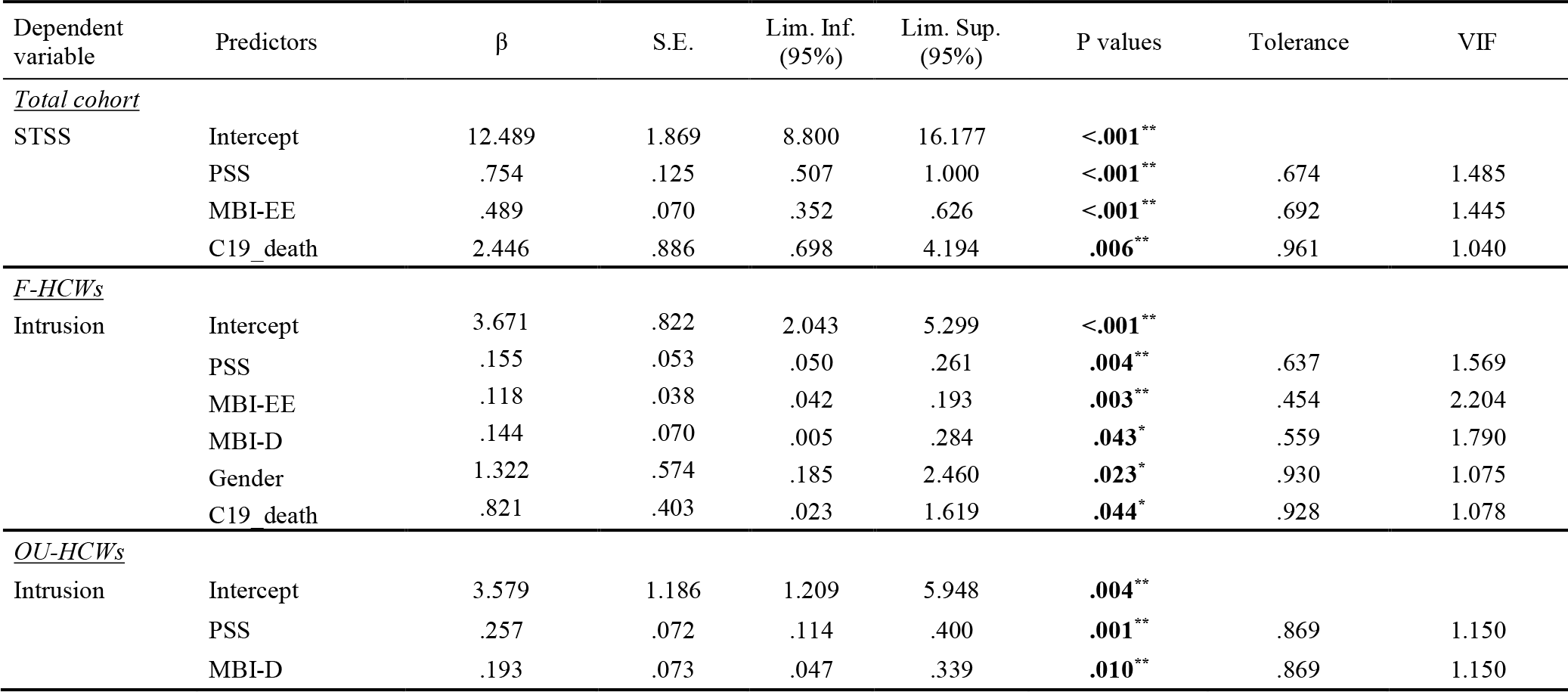
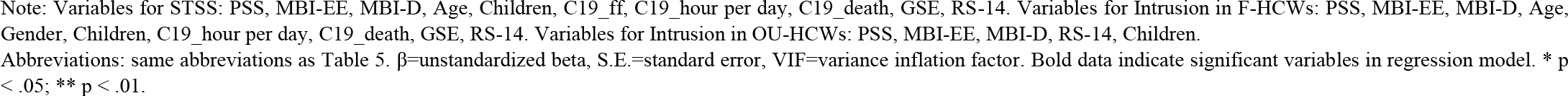
Results of stepwise multiple linear regression analysis predicting secondary traumatic stress (STSS) in total cohort (n=184) and Intrusion in F-HCWs (n=118) vs OU-HCWs (n=66).

## DISCUSSION

The aim of the present study was to assess psychological distress in terms of secondary traumatic stress and professional burnout in HCWs during the COVID-19 outbreak. A prevalence of secondary traumatization among HCWs ranging from 4% to 13% was described in studies before COVID-19 outbreak^9^, and during COVID-19 outbreak a considerable proportion of HCWs experienced mood and sleep disturbances^20,21^. We found symptoms of moderate to severe secondary traumatization in a higher proportion of the total respondents, arising over 40%. Therefore, the actual HCWs situation appears to be critical, with a prevalence even higher in frontline HCWs (47.5%) and in HCWs exposed to infected patients’ death (67.1%).

Secondary traumatic stress was positively associated with the amount of time spent with COVID-19 patients, with the exposure to COVID-19 patients’ deaths and with the severity of symptoms of family members or friends infected by COVID-19. A significant regression model was obtained and STS was positively predicted by perceived stress, emotional exhaustion, and exposure to patients’ death, confirming the central role that the unsuccessful care-taking efforts have in the development of secondary traumatization. In frontline HCWs the relationship between STS, specifically intrusion symptoms, and exposure to patients’ death as predictors was confirmed, meanwhile, it was not observed in HCWs working in other units. No significant protective factors were found. In light of these findings, we reasonably hypothesize that the observed high level of STS is consistent with the actual outbreak and therefore it’s potential long terms consequences should be taken into account.

The prevalence of professional burnout is similar to previous findings and is over 50%^26,27^. No significant differences in frontline HCWs or in HCWs exposed to patients’ death were found for the prevalence of professional burnout, suggesting that it is not so closely related to the COVID-19 outbreak. In fact, even if in our study professional burnout correlates with secondary traumatization, that’s may be due to the partial overlapping of constructs^9^. Bellolio et al.^45^ underlined that burnout is a result of the mismatch between the nature of the job and the nature of the person who does the job, it’s gradual and it arises from daily life, through continuous negative experience, without a necessarily traumatic character ^26,45,46^. Secondary traumatic stress is instead an acute reaction, secondary to relationship, that arises when rescue care-taking efforts are unsuccessful^45^. Our findings appear to be in accordance with this distinction.

In our study, frontline HCWs had significant higher scores on secondary traumatic stress, intrusion subscale, meanwhile, they exhibited a significant lower score on burnout, depersonalization subscale, when compared to HCWs involved in other units. This may be due to the fact that intrusion symptoms are characteristics of traumatic stress reactions, that particularly emerged during the COVID-19 outbreak in frontline HCWs, and may be distinctive from other symptomatology such as professional burnout. Further investigations are required. It is also possible that, in our respondents, high levels of burnout and perceived stress were already present before the COVID-19 outbreak, as similar levels were reported in HCWs in previous studies ^26,27^. This point needs to be clarified with longitudinal studies.

Our findings suggest that the COVID-19 outbreak had an impact on the more frequent direct exposure to the patients’ physical pain, psychological suffering, and death, which increased secondary traumatization in HCWs. Further investigations are required, to better clarify the longitudinal course of the effects of traumatization and the occurrence of long-term pathologic consequences. The high prevalence of STS symptoms we found in this study point out that large scale screening and treatment programs for secondary traumatic stress in HCWs, particularly to prevent long term consequences of COVID-19 outbreak are necessary.

## Data Availability

The raw data supporting the conclusions of this manuscript will be made available by the authors, without undue reservation, to any qualified researcher.

## Limitations

The present study has some limitations that should be considered. The complexity of the survey and the time required to fill the questionnaires limited the number of participants, moreover in a period in which the workload was overwhelming and the main part of respondents was directly involved with patients. The limited number of participants, make it difficult to generalize the results to the whole HCWs population. The cross-sectional nature of the study and the lack of longitudinal follow-up, do not allow inferences about the causal relations among the variables, and the long-term consequences of the psychological outcomes found. Further prospective studies could clarify the relation between burnout, secondary traumatic stress, and protective factors. Long-term implications of HCWs mental health and consequences of personal and organizational factors are worth further investigation.

## Acknowledgements

We would like to thanks for contributing to the diffusion and realization of the Survey: Hilary Pinnock^1^ and Ioannis Vogiatzis^2^ on behalf of Assembly 1 Clinical of European Respiratory Society (ERS); Antonio M Esquinas^3.^

## Footnotes

1 Univerity of Edinburgh, Edinburgh, UK;

2 Northumbria University, Newcastle upon Tyne, UK;

3 Hospital Morales Meseguer, Murcia, Spain.

